# Current Evidence of the Pharmacological Treatments for Novel Coronavirus Disease 2019 (COVID-19): A Scoping Review

**DOI:** 10.1101/2020.05.12.20093997

**Authors:** Noyuri Yamaji, Sachiko Ohde, Kimi Estela Kobayashi-Cuya, Shota Saito, Osamu Takahashi

## Abstract

**Background:** As of May 2 2020, 3,267,184 confirmed cases of COVID-19 and 229,971 COVID-19-caused deaths have been reported worldwide. Currently, there is limited clarity on the pharmacological treatments available for the novel coronavirus. We systematically identified the current evidence and ongoing research on the pharmacological treatments for COVID-19.

**Methods:** We conducted a scoping review using PRISMA-ScR. Observational studies, including cohort studies and case series, as well as experimental studies, including randomized controlled trials (RCTs) and non-RCTs were searched electronically on April 7, 2020 and by hand on May 1, 2020. PubMed, EMBASE, and Cochrane library databases were searched along with seven trial registries. The inclusion criteria were patients with confirmed COVID-19 who received pharmacological therapies, including hydroxychloroquine and chloroquine, lopinavir/ritonavir, remdesivir, tocilizumab, and favipiravir.

**Results:** We identified 222 studies on pharmacological treatment of the novel coronavirus. We included 11 of these studies in this review, including the ones on hydroxychloroquine and chloroquine (one cohort), lopinavir/ritonavir (one RCT, three cohorts, and two case series), remdesivir (one RCT and one case series), tocilizumab (one case series), and favipiravir (one RCT). In the three RCTs carried out in China, both lopinavir/ritonavir and remdesivir did not show any significant earlier clinical improvement in case of severe infection [Hazard ratio (HR): 1.31, p=0.09 and HR: 1.24, p=0.24, respectively], The clinical recovery rate on day seven was not significantly different between the favipiravir and arbidol groups (p=0.14) for moderate patients, although the duration of pyrexia and cough in the favipiravir group was significantly shorter as compared to the arbidol group (p<0.01). There are 135 ongoing RCTs, including 72 for hydroxychloroquine and chloroquine, 29 for lopinavir/ritonavir, 14 for remdesivir, 16 for tocilizumab, and 4 for favipiravir.

**Conclusion:** The clinical effectiveness and safety of these drugs for the treatment of COVID-19 remains unclear owing to the lack of large, high-quality RCTs. However, in the event of emerging infectious diseases, we need to repeatedly and systematically update the best available evidence to avoid misleading information.

## Background

The novel coronavirus disease 2019 (COVID-19), first identified in Wuhan, China, in December 2019, is a zoonotic disease caused by the family of single-stranded RNA coronaviruses. There are six recognized types of coronaviruses that can cause infections in humans, including the Middle East Respiratory Syndrome and Severe Acute Respiratory Syndrome [1]. The Coronavirus Research Group of the International Commission on Virus Taxonomy has proposed that the novel coronavirus be referred to as Severe Acute Respiratory Syndrome Coronavirus 2 (SARS-CoV-2) [2].

COVID-19 is spreading exponentially around the world and has been declared as a global pandemic by the World Health Organization (WHO) on March 11, 2020 [3]. As of May 2, 2020, 3,267,184 COVID-19 cases have been confirmed and 229,971 people have died all over the world owing to the disease [4]. COVID-19 produces an acute viral infection and causes Severe Acute Respiratory Syndrome [5,6]. Older adults, men, smokers, and people with other comorbidities are more likely to suffer from the COVID-19 infection [7].

Currently, there is no specific approved treatment for COVID-19. Various potential drugs, such as hydroxychloroquine (HCQ) and chloroquine (CQ), lopinavir/ritonavir (LPV/r), remdesivir, tocilizumab (TCZ), and favipiravir are being tested as pharmacological treatments for COVID-19 [8-10]. However, there is limited and inconclusive evidence of the clinical effectiveness of these pharmacological therapeutics [11].

## Methods

### Aim

We aimed to systematically review the positive as well as adverse effects of pharmacological treatments for COVID-19 infection, based on the current published evidence and ongoing studies.

### Study design

We performed a scoping review of pharmacological treatments for COVID-19 infection and synthesized the positive and adverse effects using PRISMA-ScR [12].

### Eligibility criteria

We included research articles on the use of five therapeutic medications of our interest for the treatment of COVID-19 infection, including HCQ and CQ, LPV/r, remdesivir, TCZ, and favipiravir. The studies consisted of randomized controlled trials (RCTs), non-RCTs, cohort studies, case control studies, and case series studies, with at least five patients each. Our search included only human studies and peer-reviewed studies published until April 7, 2020. We excluded any reviews, letters, expert opinions or research reports with incomplete information.

### Search strategy

We searched 3 databases, including PubMed, Embase, and Cochrane library, and also performed hand-searching; the keywords used were ‘COVID-19’ and ‘drug names’ (Appendix). We also searched seven clinical trial registries, including U.S. National Library of Medicine, ClinicalTrials.gov, International Clinical Trials Registry Platform, EU Clinical Trials Register, Australian New Zealand Clinical Trials Registry, International Standard Randomized Controlled Trial Number Registry, Netherlands Trial Register, University Hospital Medical Information Network while restricting our search to only intervention studies. Articles with the status ‘withdrawn’ or ‘unknown’ were excluded.

### Reference selection and data extraction

After removing duplications, two reviewers (N.Y. and S.O.) selected relevant articles by reading full-texts; one reviewer (N.Y. or K.K.) synthesized the information and constructed a summary table of the findings; and the other three reviewers (N.Y., K.K., and S.O.) confirmed all data. Two reviewers (N.Y. or S.O.) confirmed the data after being extracted by one reviewer (S.S.).

### Evaluation of study design and data synthesis

We assessed the study design of each study included in this review following the 2009 Oxford CEBM Levels of Evidence [13].

### Patient and Public Involvement

Patients and the public were not involved in this research.

## Results

We found 5,936 studies on COVID-19 and of these, 222 studies that were focused on the use of the five drugs of our interest were identified through database searching. Cochrane library had only clinical trials; thus, we included other relevant studies from PubMed and Embase as well (Figure 1). The final selection of studies included three RCTs [14-16], four cohort studies [17-20], and four case series [21-24] that were carried out in China (eight studies), Singapore (one study), France (one study) along with an international study. Drug-wise distribution of the studies was as follows: HCQ and CQ (one cohort)^17^, LPV/r (one RCT, three cohort, and two case series) [15,18-22], remdesivir (one RCT and one case series) [16,24], TCZ (one case series) [23], and favipiravir (one RCT) [14], while two of the studies were pre-proof articles (Table 1) [14,17]. We also identified 135 ongoing RCTs: 72 for HCQ and CQ, 29 for LPV/r, 14 for remdesivir, 16 for TCZ, and four for favipiravir (Figure 2). The highest number of ongoing RCTs was being carried out internationally (20), followed by Spain (18), France (17), China (15), Iran (10), and the United States (9) (Figure 3).

**Figure 1.**
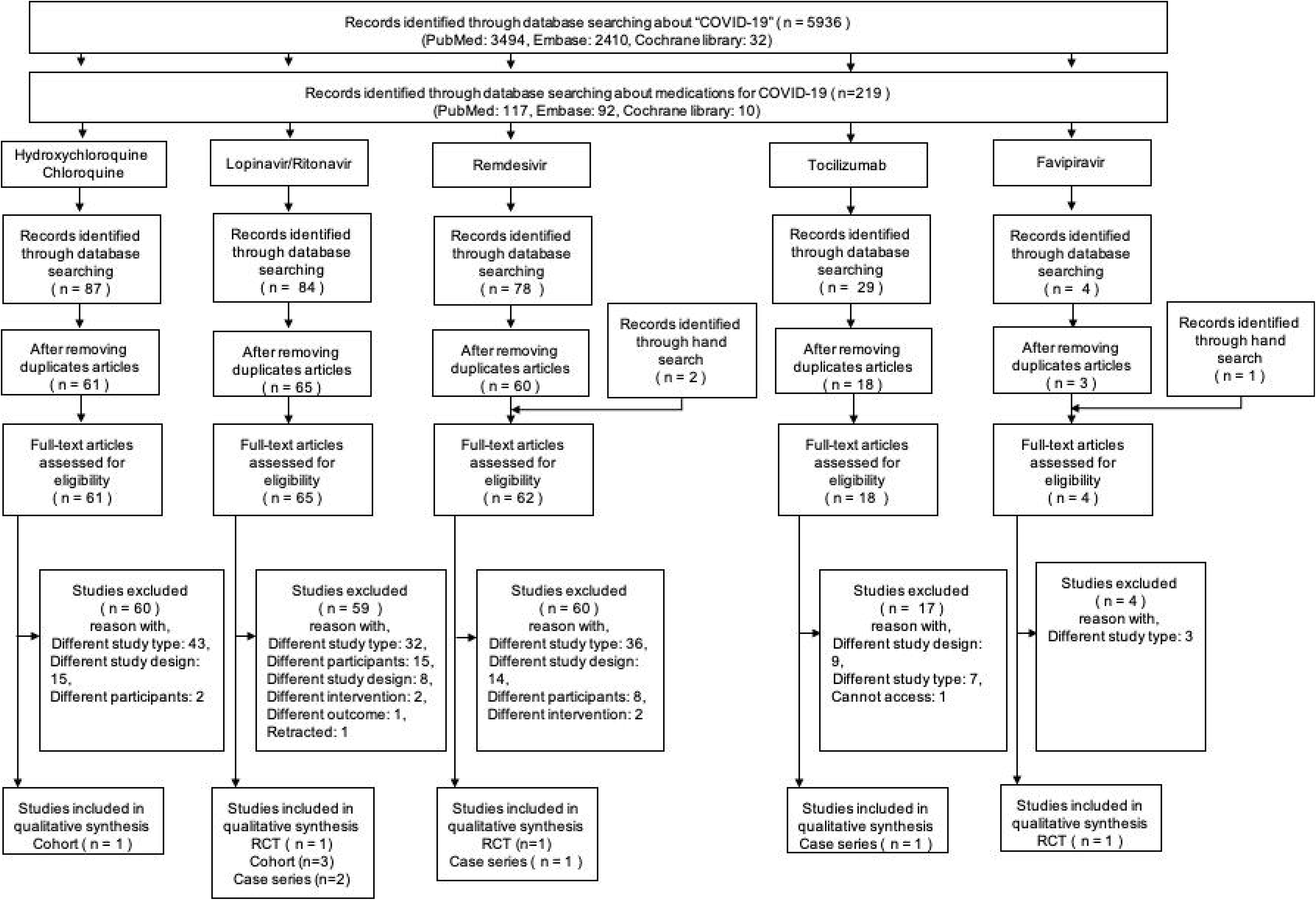
Process of screening eligible studies

**Figure 2.**
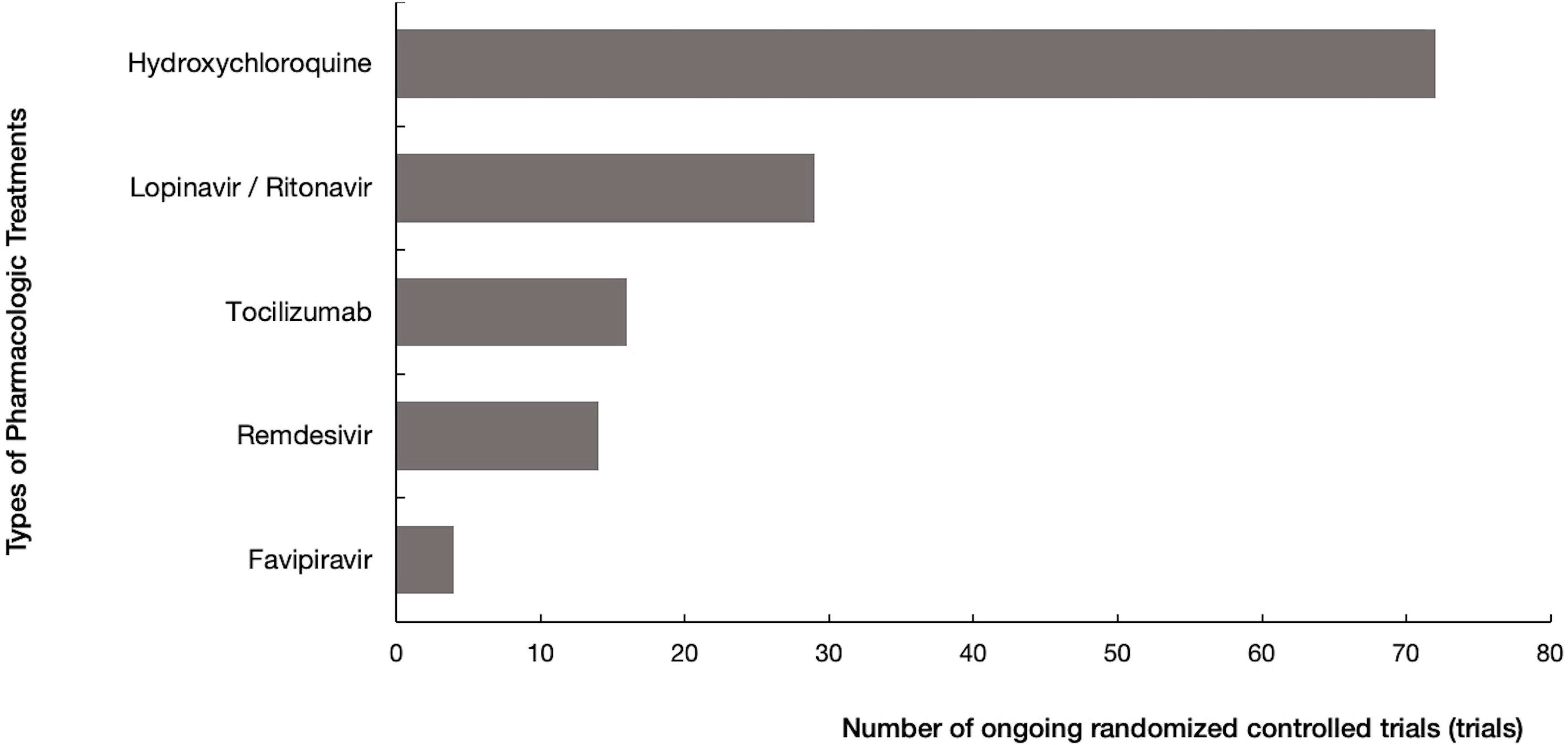
Number of worldwide registered randomized controlled trials on pharmacological treatments available for COVID-19, segregated by drug

**Figure 3.**
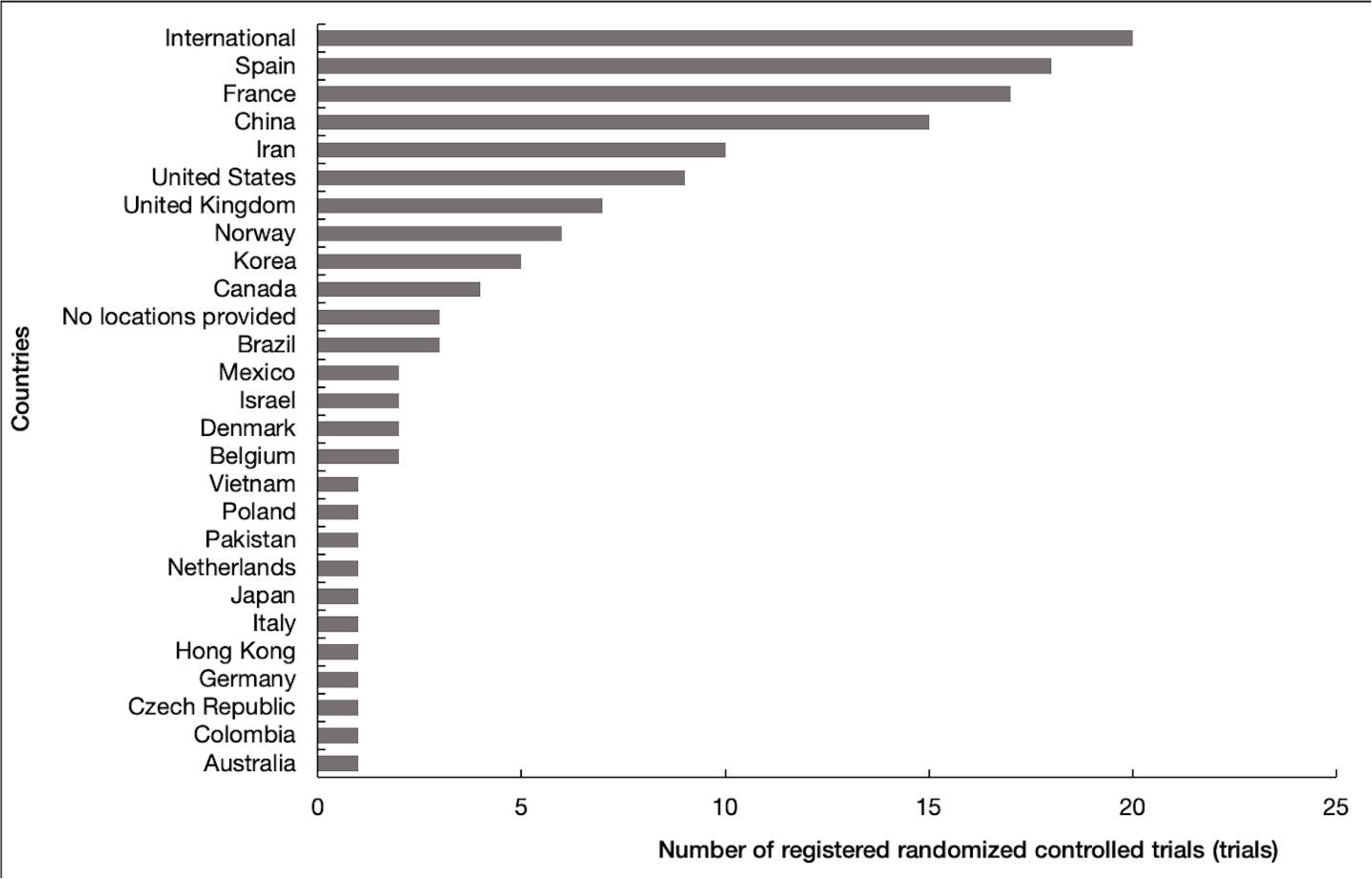
Number of worldwide registered randomized controlled trials on pharmacological treatments available for COVID-19, segregated by country

(Table 1 near here)

### Hydroxychloroquine and Chloroquine

HCQ and CQ are used for treating malaria and autoimmune diseases. Of the two, CQ was the first to be used for prevention and treatment of malaria, while HCQ has fewer side effects as compared to CQ [25]. We screened 61 articles after removing duplicates. Of the 61 articles, one cohort study was identified [17]. Gautret *et al*. conducted a prospective cohort study on 42 patients with COVID-19. Patients were allocated to the HCQ group or to the standard care group. On day 6, the proportion of viral clearance was higher in the HCQ group, as compared to the control group (HCQ group: 14/20, control group: 2/16, p<0.001) [17].

### Lopinavir/Ritonavir

LPV/r are anti-retroviral protease inhibitors that are used in combination to suppress the production and proliferation of human immunodeficiency virus [26]. We screened 65 articles after removing duplicates. Of the 65 articles, 59 studies were excluded and 1 RCT, 3 cohort studies, and 2 case series were included for this review [15,18-22]. Cao *et al*. conducted an open-label RCT with 199 hospitalized COVID-19 patients. Patients were randomly assigned to receive LPV/r along with standard care (99 patients) or standard care alone (100 patients) for 14 days. LPV/r treatment did not show any significant earlier clinical improvement (LPV/r group: median 16 days, control group: median 16 days, hazard ratio [95% CI]: 1.31 [0.95-1.85], p=0.09). The 28-day mortality in the LPV/r group was also not significantly lower than in the control group (LPV/r group: 19.2%, control group: 25.0%, rate differences [95% CI]: -5.8 [-17.3 to 5.7]). 46 (48.4%) patients in the LPV/r group and 49 (49.5%) in the control group had adverse events, while 19 (19%) in the LPV/r group and 32 (32%) in the control group had severe adverse events [15].

Ye *et al*. conducted a prospective cohort study of 47 hospitalized patients infected with COVID-19. The individuals were divided into LPV/r (LPV/r combined with pneumonia-associated adjuvant drugs) and control (only adjuvant drugs) groups. Those in the LPV/r group showed a faster decrease in body temperature, reduced levels of white blood cells, lymphocytes, and C-reactive protein (CRP) as well as quicker nCoV-RNA clearance, as compared to the control group [19].

Deng *et al*. conducted a retrospective cohort study. It included two groups, a combination group that received oral arbidol and LPV/r and a monotherapy group that received only oral LPV/r after diagnosis. After two weeks of treatment, 94% and 53% patients in the combination and monotherapy groups, respectively, tested negative for SARS-CoV-2. Moreover, after one week of treatment, 69% and 29% patients in the combination and monotherapy groups, respectively, showed a significant improvement in chest CT scans [18].

Yuan *et al*. conducted a retrospective cohort study with 94 discharged patients who displayed mild, moderate, and severe symptoms of COVID-19 infection. COVID-19 mRNA clearance ratio, as detected by PCR, was found to be significantly correlated with the length of hospitalization in patients treated with IFN-α+LPV/r or IFN-α+LPV/r+ribavirin [20].

Wan *et al*. conducted a case series of 135 hospitalized COVID-19 patients. All patients received LPV/r and interferon as anti-viral therapy. A total of 15 patients (11%) were discharged, while one patient died [21]. Young *et al*. conducted a case series on 18 hospitalized patients diagnosed with COVID-19. LPV/r was prescribed to five patients who suffered from oxygen desaturation and required supplemental oxygen. Of these, three patients showed a reduction in their oxygen requirements within three days of taking the drug, while the viral shedding from the nasopharyngeal swabs of two patients disappeared within two days. Contrarily, two of the five patients showed continued viral shedding and progressive respiratory failure, with one of them requiring invasive mechanical ventilation [21].

### Remdesivir

Remdesivir is a novel anti-viral drug developed by Gilead Sciences that is used for the treatment of Ebola virus disease. It is not yet licensed or approved anywhere globally for the potential treatment of COVID-19 [27]. We screened 61 articles after removing duplicates. Of the 61 articles, one RCT and one case series were included in this study [16,24].

Wang *et al*. conducted a double-blind RCT with 237 COVID-19 patients. All the patients had an oxygen saturation of 94% or less and confirmed pneumonia. They were randomly assigned to either the remdesivir group or the placebo group and received the treatment for 10 days. No significant differences in terms of clinical improvement, hospitalization length, and 28-day mortality were observed between the two groups. Furthermore, due to adverse events, more patients in the remdesivir group (12%) discontinued the study as compared to the placebo group (5%) [16].

Grein *et al*. conducted a case series of 53 hospitalized patients (from 9 different countries) diagnosed with COVID-19 using reverse-transcriptase polymerase chain reaction (RT-PCR) assay. Patients who were receiving oxygen support or who had an oxygen saturation of 94% or less received remdesivir for 10 days. After receiving the first dose of remdesivir, 36 of the 53 patients showed an improvement in the need for oxygen support. During a median follow-up of 18 days, 17 of the 30 patients who were receiving mechanical ventilation were extubated, while extracorporeal membrane oxygenation was stopped for three of the four patients who were receiving it. A total of 25 patients were discharged, and eight patients got worse, of which, seven died at least 28 days after taking remdesivir [24].

### Tocilizumab (TCZ)

TCZ is a recombinant humanized monoclonal antibody that acts as an interleukin (IL)-6 receptor antagonist and is used in the treatment of rheumatism [28]. We screened 18 articles after removing duplicate studies. Of the 18 articles, one case series was included [23]. Luo *et al*. conducted a case series on 15 hospitalized patients diagnosed with COVID-19. Patients received 80-600 mg TCZ each time; 8 of them received TCZ with methylprednisolone. The CRP levels of 11 patients returned to the normal range within one week, while three of the four critically ill patients who received a single dose of TCZ died; one of them did not return to normal CRP levels within a week [23].

### Favipiravir

Favipiravir is a novel anti-viral agent that selectively inhibits RNA-dependent RNA polymerase of RNA viruses. It is prescribed for patients only when other anti-influenza virus drugs are ineffective or are insufficiently effective in the event of re-occurrence of the influenza virus infection or when there is no known cure for an RNA virus infection [29]. We screened four articles after removing duplicates and one pre-proof RCT met our inclusion criteria [14].

Chen *et al*. conducted an open-label multicenter RCT on 240 hospitalized patients diagnosed with COVID-19 pneumonia. Patients were randomly assigned to either the umifenovir (arbidol) or favipiravir groups. Clinical recovery rate (recovery of body temperature, respiratory rate, oxygen saturation, and cough relief) on day 7 was not significantly different between both the groups (favipiravir group: 71/116, arbidol group: 61/120, p=0.14, Difference of recovery rate: 0.0954 [95% confidence interval: -0.0305-0.2213]). The duration of pyrexia and cough in the favipiravir group was significantly shorter than in the arbidol group (pyrexia, difference: 1.70 days (p<0.01) and cough, difference: 1.75 days (p<0.01), respectively). A post-hoc analysis showed that the incidence of de *novo* dyspnea in the favipiravir group was significantly lesser than in the arbidol group (favipiravir group: 4/116, arbidol group: 14/120, p<0.0174). 37 patients in the favipiravir group and 28 patients in the arbidol group had level 1 adverse effects [14].

## Discussion

In this scoping review that aimed to systematically clarify the evidence for the use of five potential drug treatments for COVID-19. Based on the 11 studies analyzed (including only three RCTs carried out in China), we found that the clinical effectiveness and safety of these pharmacological drugs remains controversial.

One cohort study showed that the HCQ-administered group displayed a higher proportion of efficacy for viral clearance, as compared to the group not administered HCQ [17]. Among the six studies related to LPV/r [15,18-22], four studies reported that LPV/r may be effective in viral shedding [18-20,22] and two studies reported that it may improve clinical conditions, such as CT results and body temperature [19,21]. However, the RCT by Cao *et al*., which included the largest population of all the included studies and set a clear comparison group, concluded that compared to the standard treatment, no apparent benefit of LPV/r was observed in patients hospitalized with severe COVID-19. Since adverse events of this medication have not been fully reported, the effectiveness of LPV/r is still debatable [15]. This suggests that the use of this medication for the treatment of COVID-19 should be handled with special care. In a case series study on remdesivir that included patients from multiple countries, although 84% of the patients showed clinical improvement, there was a high adverse event rate (32/53); these events included increased hepatic enzymes, diarrhea, rash, renal impairment, and hypotension [24]. On the contrary, one RCT carried out in China showed no significant difference between the remdesivir group and the placebo group in terms of clinical improvement, hospitalization length, and mortality. Furthermore, more patients in the remdesivir group (12%) discontinued the medication due to its adverse effects [16]. To confirm its effectiveness, further high-quality RCTs need to be conducted. Furthermore, in a case series study using TCZ on 15 patients, an improvement was observed in the levels of CRP and interleukin-6 receptor (IL-6R). Moreover, neither clinical conditions nor deaths were reported following this medication [23]. When favipiravir was compared with arbidol, no clear difference in the clinical recovery rate was observed between the two groups on day 7, although the duration of pyrexia and cough in the favipiravir group was found to be significantly shorter than in the arbidol group [14]. Considering that we identified only a limited number of studies reporting on the effectiveness of pharmacological treatments during the course of conducting this review, the results presented here should be treated with caution.

Although the primary endpoints for most of the studies included were within two weeks of dosage, symptoms were found to persist beyond this time period as well. For instance, three studies were followed up for at least 28 days [15-16,24], but further trials should have been conducted for a longer time. Also, in two open-label RCTs, personal and outcome assessments should have been blinded to avoid performance and detection biases [30]. It is essential for a systematic review to be conducted by collecting data from high-quality RCTs in order to provide the best evidence [13]. Moreover, it needs to be free of heterogeneity in the direction and degree of the results between RCTs [13]. In this regard, to meet the optimal information size criteria, RCTs require to be conducted with larger sample sizes (at least between 2,000-4,000 patients) to yield low imprecision results with a narrow 95% confidence interval [31]. In addition, large, high-quality RCTs are needed to determine the potential positive as well as adverse effects of pharmacological treatments for COVID-19.

In total, we found 135 ongoing RCTs, including 72 for HCQ and CQ, 29 for LPV/r, 14 for remdesivir, 16 for TCZ, and four for favipiravir. While these RCTs would help improve the certainty of the evidence, interpretation of their findings need to be handled with care, especially when these are sponsored by pharmaceutical companies, as such studies may have a higher possibility of substantial bias [32].

In addition to the drugs that we searched for in this study, some institutions have announced the development of new drugs for COVID-19. For example, Biotest, BPL, LFB, and Octapharma have joined an alliance formed by CSL Behring (ASX:CSL/USOTC:CSLLY) and are collaborating with Takeda Pharmaceutical Company to develop a potential plasma-derived therapy for treating COVID-19 [33]. REGENERON is developing a new cocktail drug of two antibodies [34]; however, the development is still in its early stage. Also, WHO reported that it is hard for people to find accurate guidance regarding the 2019-nCoV outbreak and response because of an ‘infodemic’, defined as an over-abundance of information [35]. For this reason, it is extremely important to update and provide the current best evidence of pharmacological treatments for COVID-19 and make a medical decision without being misled by inappropriate information.

## Limitations

This review has some limitations. First, we included data studies published until April 7, 2020; however, since COVID-19 is a rapidly emerging infectious disease, new research on it is being published on a daily basis. There are several RCTs that are currently in progress, which will help improve our limited knowledge and clarify the effects of these treatments. A systematic review of the pharmacological treatments for COVID-19 infection, including RCT results, would clarify their effects. Tricco *et al*. suggest that a scoping review protocol is necessary to ensure transparency and reduce duplication of effort [12]. Although we were not able to submit the research protocol, it is important to note that in case of public health emergencies such as COVID-19, it is crucial to provide the current related-information as soon as possible. In the future, we need to register and carry out the protocol according to the research plan when conducting a systematic review for each of these drugs.

## Conclusions

This scoping review identified that the clinical effectiveness and safety of the currently available pharmacological treatments for COVID-19 remain unclear due to the lack of large, high-quality RCTs. In the event of emerging infectious diseases such as COVID-19, we need to systematically update the best available evidence being generated by ongoing international RCTs to avoid misleading information.

## Data Availability

All data analyzed during this study are included in this published article (and its supplementary information files).

## List of abbreviations

RCT: randomized control trial
WHO: World Health Organization
HCQ: hydroxychloroquine
CQ: chloroquine
LPV/r: lopinavir/ritonavir
TCZ: tocilizumab
HR: hazard ratio
CRP: C-reactive protein

## Declarations

### Ethics approval and consent to participate

Not applicable.

## Consent for publication

Not applicable.

## Availability of data and materials

Not applicable.

## Competing interests

No declaration.

## Funding

This research received no specific grant from any funding agency in the public, commercial or not-for-profit sectors.

## Author contributions

NY and OT were responsible for the conception and design of the study. NY, SO, KK, and SS were responsible for the acquisition of data and analysis. NY, SO, and KK wrote the manuscript. All the authors participated in the critical revision of the manuscript for important intellectual content. All the authors had full access to all the data in the study and take responsibility for the integrity of the data and the data synthesis. SO and OT supervised the conduction of this review

## Acknowledgements

Prof. Erika Ota, PhD (Representative Director of Japan Branch of the Cochrane Collaboration and St. Luke’s International University) provided important insights to the methodology of this manuscript. We would like to thank Editage (www.editage.com) for English language editing.

## Notes

### Competing Interest Statement

The authors have declared no competing interest.

